# Publication bias in otorhinolaryngology meta-analyses in 2021

**DOI:** 10.1101/2023.07.24.23293129

**Authors:** Fatemeh Mohammadian, Shahin Bastaninejad, Shirin Irani

## Abstract

**Introduction:** One concern in meta-analyses is the presence of publication bias (PB) which leads to the dissemination of inflated results. In this study, we assessed how much the meta-analyses in the field of otorhinolaryngology in 2021 evaluated the presence of PB.

**Methods:** Six of the most influential journals in the field were selected. A search was conducted, and data were extracted from the included studies. In cases where PB was not assessed by the authors, we evaluated the risk of its presence by designing funnel plots and performing statistical tests.

**Results:** 75 systematic reviews were included. 51% of them used at least one method for assessing the risk of PB, with the visual inspection of a funnel plot being the most frequent method used. 29% of the studies reported a high risk of PB presence. We replicated the results of 11 meta-analyses that did not assess the risk of PB and found that 63.6% were at high risk.

**Discussion:** Our results indicate that systematic reviews published in some of the most influential journals in the field do not implement enough measures in their search strategies to reduce the risk of PB, nor do they assess the risk of its presence.

## 1 Background

### 1-1 Rationale

The Catalogue of Bias, a collaboration dedicated to describing a wide range of biases and outlining their potential impact on research studies, defines publication bias (PB) as “when the likelihood of a study being published is affected by the findings of the study”^1^. Several factors could lead to PB including selective publication of positive findings, selective publication of statistically significant findings, selective publication of ‘interesting’ findings, and publication according to the quality of the trial or its funding ^2^. A study in 2009 demonstrated that randomized controlled trials with positive findings were 3.9 times more likely to be submitted and published than trials with negative or null findings ^3^. The OPEN project (Overcome failure to Publish nEgative fiNdings), a project funded by the European Union to investigate the extent and impact of dissemination bias, has encouraged systematic reviewers (SRs) to follow “the best practices” in conducting systematic reviews (SR) (especially practices concerning the assessment of the impact of dissemination bias) and publishing the protocol and results of their SRs publicly. Regarding “the best practices” in conducting SRs, the members of the OPEN project have proposed the Cochrane Handbook for Systematic Reviews of Interventions ^4^ and the standards for SRs stated by the Institute of Medicine of the National Academy of Sciences ^5^. The main recommendation in these guidelines for avoiding PB is to search for evidence that includes bibliographic databases and other sources such as grey literature, citation indexes, trial data, and other unpublished reports. On the other hand, selection models and graph-based methods have been suggested to be the main methods for assessing the presence of PB ^6^.

Unfortunately, despite the significance of PB, previous studies show that a considerable proportion of systematic reviewers in different fields, such as oncology ^7^, anesthesiology ^8^, dermatology ^9^, cardiology ^10^, and gastroenterology ^11^ did not try to evaluate its possible presence in their SRs and MAs. Also, a substantial proportion of SRs were found not to search resources other than published materials, hence increasing the risk of PB in their results ^12^.

In this study, we aimed to assess to what extent the recent SRs with MAs in the field of otorhinolaryngology (ENT) have taken measures to reduce the risk of PB in their results or have evaluated the probability of its presence in their research. To our knowledge, this is the first study that has evaluated this subject in this field. Our findings may help to understand how much the issue of PB is addressed in the field.

### 1-2 Approaches to deal with publication bias

There are two approaches for dealing with PB: selection models and graph-based methods ^6^. Selection models use the weighted distribution theory to model the publication process and thus, develop estimation procedures that account for the selection process ^13^. The Hedges model ^14^ is the first and most known selection model used for assessing PB. Unfortunately, these models rely on largely untestable assumptions ^15^, and thus, are rarely used in practice for assessing PB. Instead, they are used in sensitivity analyses ^13^. On the other hand, graph-based methods are widely used. These methods are based on a funnel plot which usually presents effect sizes plotted against their standard errors (SE) or precision ^16^. In the presence of PB, the plot is expected to be asymmetrical. However, a variety of other factors may also lead to asymmetrical funnel plots, such as inflated effects in smaller studies, true heterogeneity, artefactual effects, and chance ^17^. Another issue with the visual inspection of funnel plots for detecting PB is the subjectivity of the approach, which in turn leads to errors in interpretation ^18^. Thus, statistical tests have also been proposed to detect funnel plot asymmetries, such as Begg’s rank test ^19^, Egger’s regression ^17^, Harbord’s regression ^20^, Peters’ regression ^21^, and Deeks’ regression ^22^. Trim-and-fill method, a nonparametric rank-based correction method, was proposed to recover symmetry by “trimming” observed studies and subsequently imputing missing studies ^23^.

It has been proposed that reviewers use various tests and methods to detect PB in their research because different tests make different assumptions on the association between the effect sizes and their precision measures ^24^. The handbook of Cochrane ^25^ has also made some other recommendations:

- tests should be used only when there are at least 10 studies included in the meta-analysis;
- tests should not be used if studies have similar SE of effect estimates;
- results of tests should be interpreted in the light of visual inspection of the funnel plot; and
- when there is evidence of plot asymmetry from a test, PB should be considered as only one of several possible explanations.

### 1-3 Objectives

To what extent the MAs in the field of ENT in 2021 have searched for evidence other than bibliographic databases and assessed the risk of PB?

## 2 Methods

### 2-1 Study selection

Six of the ten journals with the highest impact factor in the field of ENT (discovered through Google Scholar) were selected following a consensus between a group of four attending ENT surgeons at the Tehran University of Medical Sciences. These included: *Audiology and Neurotology*, *Ear & Hearing*, *International Forum of Allergy & Rhinology*, *Otolaryngology-Head and Neck Surgery*, *Rhinology*, and *The Laryngoscope*. PubMed was searched for the papers published in these journals in the year 2021. The “Systematic Review” and “Meta-Analysis” filters of PubMed were activated. The results of the search were imported into the Mendeley Reference Manager application, a software designed for the management of citations. Then, using the “Look up metadata by DOI” feature of the application, all the metadata of the records were updated.

Two reviewers retrieved the full texts of the records and independently assessed them for eligibility. Discrepancies were resolved through discussion. Studies that had an MA component were included.

### 2-2 Data items

Two reviewers independently extracted the data from eligible studies. Discrepancies were resolved through discussion. The following data were gathered:

- study characteristics;
- reporting guideline;
- bibliographic databases and citation indexes searched;
- other sources of data (clinical trials registries, grey literature, citation checking, etc.);
- whether studies in a language other than English were considered eligible for inclusion;
- the number of studies included in the SR;
- if assessed, the method used to assess PB;
- number of papers that cited the SR; and
- review type.

The review type was determined according to the 10 categories proposed by a typology study ^26^: effectiveness, experiential, costs/economic evaluation, prevalence and/or incidence, diagnostic test accuracy (DTA), etiology and/or risk, expert opinion/policy, psychometric, prognostic, and methodology. In cases where one SR fell into more than one category, we considered the main objective of the SR to determine the review type.

### 2-3 Publication bias assessment

We recorded the methods used for assessing PB in each SR. For SRs that did not evaluate the risk of PB, we assessed it by replicating the results of their Mas (using the reported effect measures for each included study), designing funnel plots, using the trim-and-fill method, and performing Begg’s rank test and Egger’s regression (or Deeks’ regression in case of DTA SRs). Only SRs that included at least 10 studies in their MA were assessed to comply with the beforementioned criteria.

We used R version 4.2 ^27^ “meta” package ^28^ to perform analyses and assess PB in SRs where it was not assessed. The significance level for PB tests was set at P<0.10.

## 3 Results

### 3-1 Study flow

The search on PubMed returned 188 records. After updating the metadata, 57 records were removed as they were not published in 2021. After screening the full-text papers, 56 reports were also excluded. Finally, 75 SRs were included in this study (check the **Appendix** for the characteristics of the included studies). **Figure 1** shows the flow of study selection.

**Figure 1.**
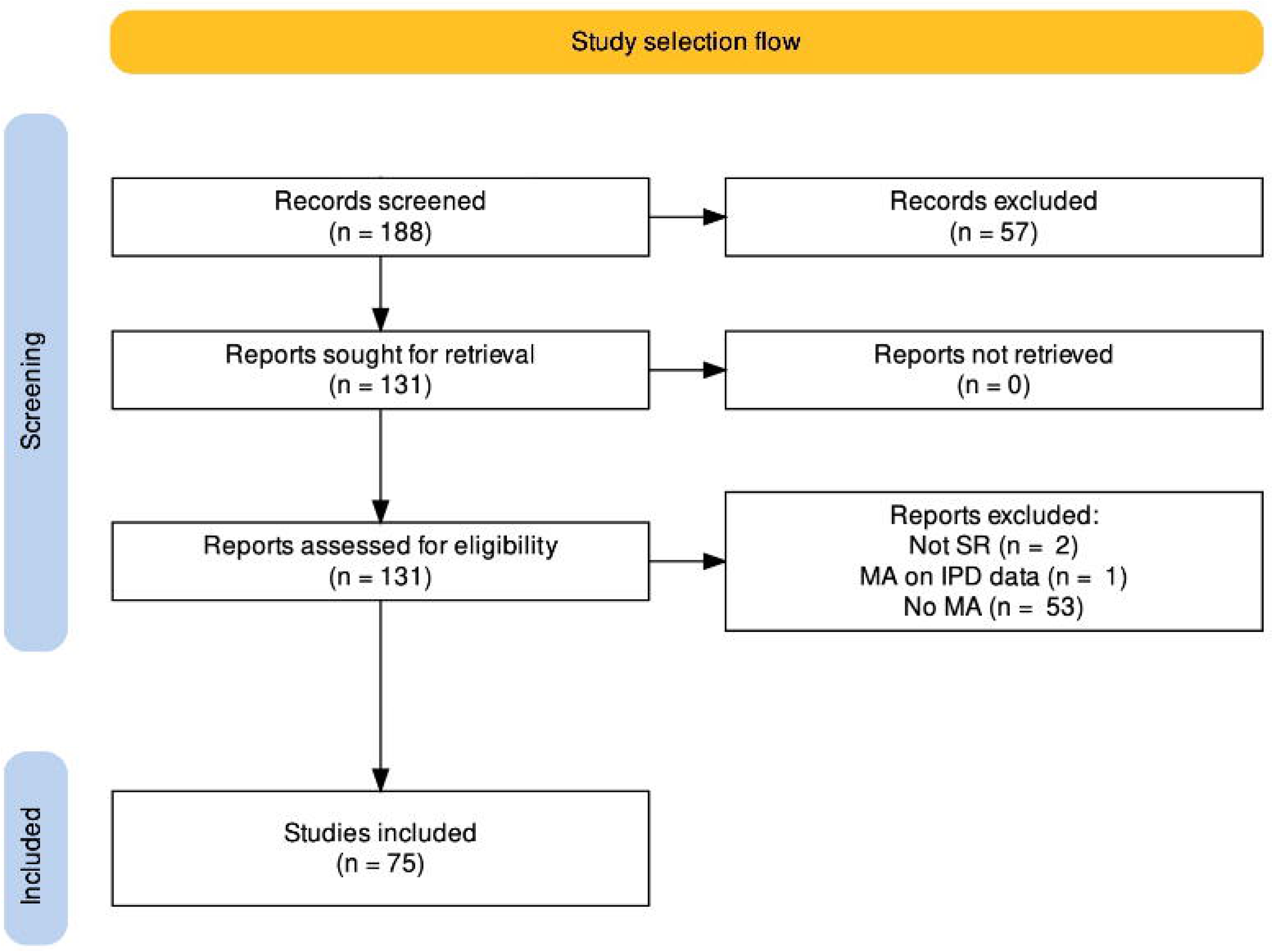
The flow of study selection.

### 3-2 Basic characteristics of contributing studies

The basic characteristics of the included SRs are presented in **Table 1**.

**Table 1.** Basic characteristics of contributing studies. SR: Systematic reviews.

| **Journal** | *Audiology and Neurotology* | *Ear & Hearing* | *International Forum of Allergy & Rhinology* | *Otolaryngology-Head and Neck Surgery* | *Rhinology journal* | *The Laryngoscope* |
| --- | --- | --- | --- | --- | --- | --- |
| **SR count** | 1 | 2 | 6 | 22 | 7 | 37 |
| **Cited by** | 0 | 27 | 15 | 208 | 16 | 394 |
| **SR type** |  | Effectiveness | Etiology and/or risk | Prevalence and/or incidence | Prognostic | Diagnostic test accuracy |
|  |  | 34 | 24 | 10 | 5 | 2 |
| **Reporting guideline** |  |  |  |  | Yes | No |
|  |  |  |  |  | 66 | 9 |

The SR types were as follows: 34 effectiveness, 24 etiology and/or risk, 10 prevalence and/or incidence, 5 prognostic, and 2 DTA. Most of the SRs followed PRISMA ^29^ or MOOSE ^30^ reporting guidelines. Only 9 SRs did not report the use of a standard reporting guideline.

Included SRs were cited by other papers on a median of 4 citations per SR (interquartile range (IQR): 1–8). The highest number of citations to an SR was 107 ^31^, while 9 SRs were not cited by any other paper at all.

### 3-3 Literature search

#### 3-3-1 Bibliographic databases and citation indexes

On average, 3.8 bibliographic databases and citation indexes were searched in the included studies. The data sources searched included APA PsychINFO, CAB Abstracts, CINAHL, CNKI, Cochrane Library, CQVIP, Embase, EmCare, Google Scholar, LILACS, MEDLINE, OTseeker, ProQuest, PubMed, SciELO, ScienceDirect, Scopus, Wanfang Data, and Web of Science. PubMed was the most searched database with 56 SRs (74.7%) searching it, followed by Embase which was searched by 51 SRs (68.0%). **Figure 2** shows how many SRs went through searching each of these databases.

**Figure 2.**
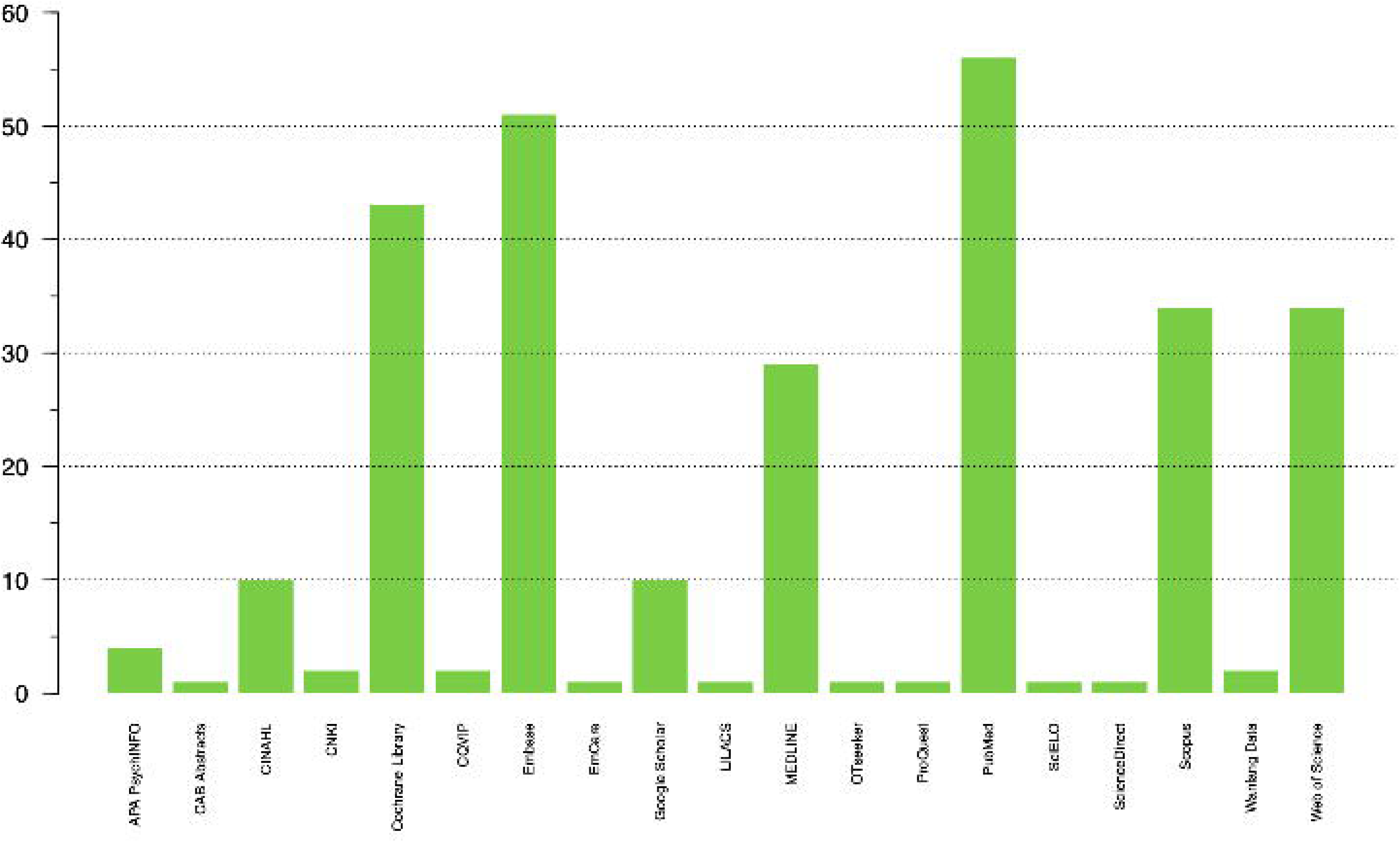
Number of included systematic reviews (SRs) that searched each bibliographic database and citation index.

#### 3-3-2 Other sources and languages

Backward citation checking was the most prevalent technique used for finding other potentially eligible records (50 SRs, 66.7%). On the other hand, forward citation checking was rarely used (4 SRs, 5.3%). Also, 3 SRs (4.0%) hand-searched the relevant journals or contacted experts in the field. Finally, 1 SR used the “related articles” feature of PubMed to search for other studies.

Search for trial registers and trial results registers was also rarely used (10 SRs, 13.3%). The registries searched were ClinicalTrials.gov (9 SRs) and ICTRP (International Clinical Trials Registry Platform) (1 SR).

The search for unpublished studies and grey literature was also less than satisfactory. One study searched for the abstracts from the related conferences, one study searched for theses (using ProQuest), one study searched the National Rehabilitation Information Center, and one study searched the Chulalongkorn Medical Library.

Most of the included SRs also used language restrictions in their search strategy. Only 18 SRs (24.0%) searched for studies in languages other than English, while 6 SRs did not report if they utilized any language restrictions in their search.

#### 3-3-3 Included studies

On average, 24.8 studies were included in the SRs (median: 16, IQR: 10.5–30.5). The maximum number of studies included in an SR was 98, while the minimum was 3. For MAs, an average number of 21.9 studies were used (median: 13, IQR: 7.5–26.5). The maximum and the minimum number of studies included in an MA was similar to the beforementioned numbers.

### 3-4 Publication bias

#### 3-4-1 Assessment for publication bias

Almost half of the included SRs used at least one method to assess the potential risk of PB (38/75, 50.7%). **Figure 3** shows the ratio of SRs that assessed PB per journal.

**Figure 3.**
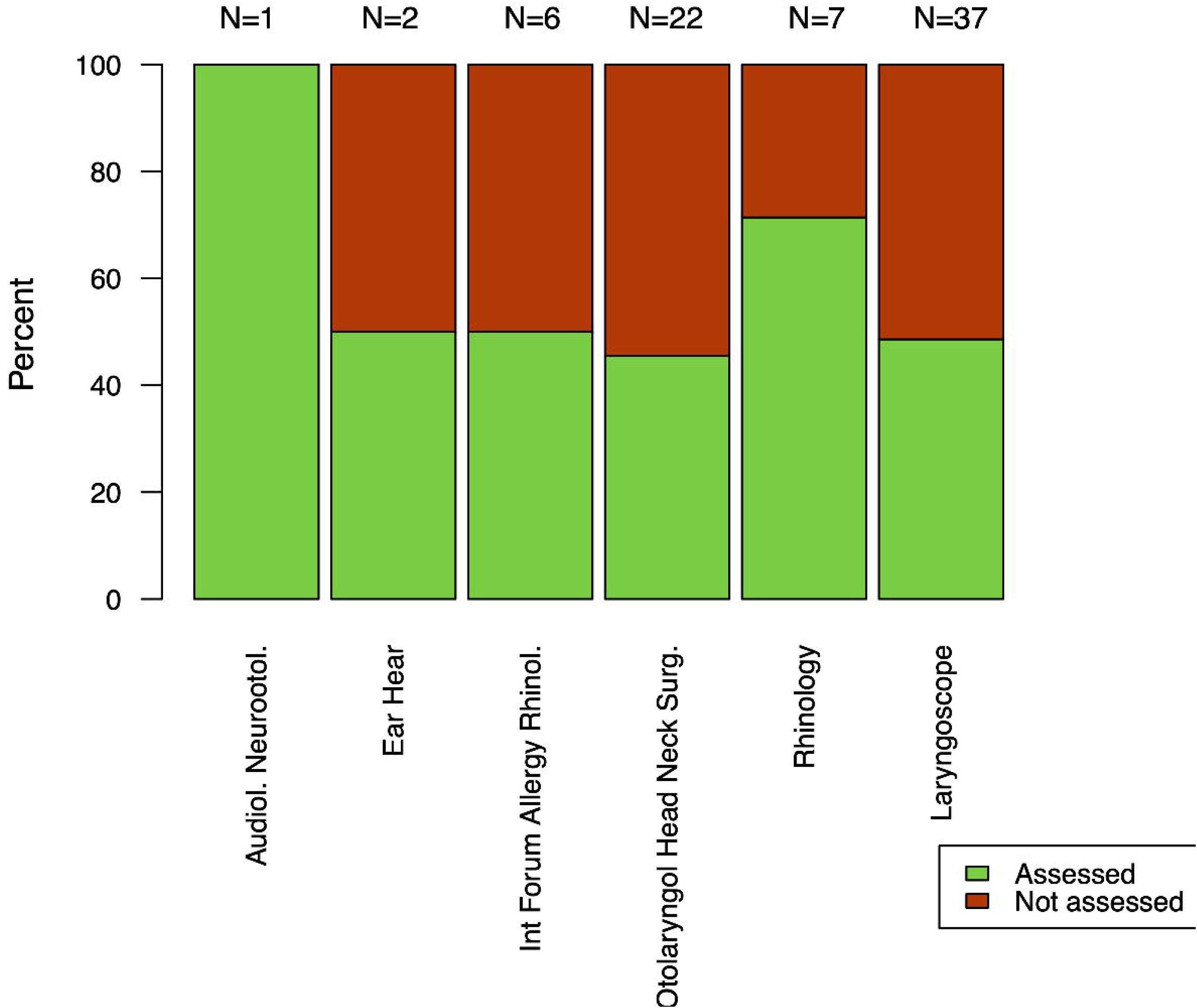
Ratio of included systematic reviews (SRs) that assessed publication bias per journal.

Visual inspection of the funnel plot was the most used method for assessing PB across the included SRs (34/38, 89.5%). Among tests used to assess funnel plot asymmetry, Egger’s regression was the most widely used (25/38, 65.8%), followed by Begg’s rank test (4/38, 10.6%). Two SRs used Harbord’s regression and one used Peters’ regression. Finally, 4 SRs used the trim-and-fill method to assess the effect of missing studies.

Of 38 SRs that assessed PB, 26 (68.4%) used at least two of the mentioned methods, while 6 (15.8%) used at least three methods. Interestingly, 4 SRs did not design a funnel plot but relied solely on statistical tests for funnel plot asymmetry to assess the potential risk of PB. Also, both included DTA SRs used Egger’s regression test, instead of Deeks’ regression, to assess funnel plot asymmetry.

#### 3-4-2 Presence of publication bias

Of the 38 SRs that assessed PB, 11 (28.9%) reported a considerable risk for its presence. We tried to assess the risk of PB in the remaining 37 SRs. In the process, we found that 22 SRs had included less than 10 studies in their MAs for any single outcome, and thus, were ineligible for PB risk assessment. Furthermore, replicating the MA for 4 SRs was impossible mostly due to an incomplete report of the data or statistical methods used. Eventually, we managed to assess the risk of PB in 11 additional SRs. Considerably, 7 of these SRs (63.6%) were found to be at a considerable risk of PB. Overall, 49 SRs were assessed for the risk of PB, out of which, 18 (36.7%) were found to be at considerable risk.

#### 3-4-3 Possible factors contributing to the risk of publication bias

To test for the possible factors that may have affected the risk of PB, a series of Pearson’s chi-square tests and t-tests were conducted. However, none of these tests were found to indicate a possible statistically significant correlation. The results of these tests are provided in **Table 2**.

**Table 2.** The possible factors which may have contributed to the risk of publication bias. OR: Odds Ratio.

| Factor |  | Risk for publication bias |  | OR | p-value |
| --- | --- | --- | --- | --- | --- |
|  |  | High risk | Low risk |  |  |
| Language restriction | Only English | 12 | 20 | 1.35 | 0.899 |
|  | Other languages too | 4 | 9 |  |  |
| Sources other than bibliographic databases (citation searching not included) | Searched | 3 | 6 | 0.83 | 0.922 |
|  | Not searched | 15 | 25 |  |  |
| Sources other than bibliographic databases (citation searching included) | Searched | 13 | 22 | 1.06 | 0.976 |
|  | Not searched | 5 | 9 |  |  |
| Number of databases searched | Mean (SD) | 3.72 (0.89) | 3.84 (1.13) | - | 0.692 |

As these results indicate, the chances of PB were slightly altered following the inclusion of languages other than English and sources other than bibliographic databases in the search strategy, but none of these correlations were statistically significant. The number of bibliographic databases and citation indexes searched was also not different in a statistically significant manner between the studies with low risk of PB vs. those at high risk of PB.

## 4 Discussion

### 4-1 Interpretation of results

In this study, we aimed to evaluate to what extent the SRs and MAs published in some of the most influential journals of ENT use methods to reduce the risk of PB and different techniques to assess the risk of its presence. Our findings revealed that this issue is not addressed optimally in a considerable proportion of the SRs.

First, the search strategies used in these SRs were not comprehensive enough to mitigate the risk of PB. Most SRs restricted their search to papers published in English, thus suffering from a great risk of language bias. Although previous studies have shown that the impact of language bias is negligible on the results of an SR in most circumstances ^32–34^, exceptions have also been observed ^35–37^. As a result, the Cochrane Initiative recommends that language restrictions should not be used unless in the setting of rapid reviews, and even in that setting, its use should be justified by the reviewers ^4^. Also, most of the SRs did not search for other sources of data or grey literature. This issue is of great importance as it has been found that such data can seriously affect the results of an SR ^38, 39^. Specifically, including a grey literature search should be seriously considered when conducting an SR because an association between “statistically significant” results and publication has been documented in previous studies ^4^.

Another finding of interest was that almost half of the SRs did not assess the risk of PB. This finding becomes bolder knowing that our analyses revealed the risk of PB was considerably higher in the SRs that did not assess the risk of PB. The reason behind this phenomenon is unknown, but some of the potential reasons could be: (a) reviewers trying not to downgrade the confidence in their results; (b) lack of methodological expertise for assessing the risk of PB which also resulted in designing poor search strategies; and (c) solely due to chance. Nevertheless, the journal editors and reviewers should ask the authors to assess the risk of PB in their SRs whenever feasible.

More importantly, we saw that in cases where reviewers found a high risk for PB in their SRs, they still did not try to expand their search. This issue should be specifically noted by journal editors, asking the authors to include other sources of data as well when the risk of PB was assessed to be high, in an attempt to avoid publishing inflated results as much as possible.

Another finding of interest was the inappropriate use of methods to assess the risk of PB. Although this problem was not frequent across the SRs, some used inappropriate tests to assess funnel plot asymmetry, such as using Egger’s regression test instead of Deeks’ regression in the setting of DTA SRs or using statistical tests alone with no visual inspection of the funnel plot beforehand. Both journal editors and reviewers must note that the results of statistical tests for funnel plot asymmetry should be interpreted in light of the visual inspection of the plot, as all these tests are known to have low statistical power ^25^. Other factors should also be considered for using such tests, such as the fact that they are not recommended for cases when there are less than 10 studies included in the MA or that they should not be used when studies are of similar size ^25^. Using contour-enhanced funnel plots is also highly desirable as they help with differentiating the reasons for funnel plot asymmetry ^40^.

Finally, we assessed some possible factors that might have contributed to the risk of PB presence. Surprisingly though, none of those factors (language restriction, a search of sources other than bibliographic databases, and the number of databases searched) had a statistically significant correlation with the presence of PB. This could be due to some possible reasons: first, it might be due to the small sample size of SRs included in the test. Another reason could be that some risk of PB was inevitable even in the absence of language restriction of the search, seeking other sources of data, and searching a large number of databases. Nevertheless, the results of these tests do not exclude the fact that implementing these measures most probably will reduce the risk of PB.

### 4-2 Implications

Our findings indicate the lack of methodological sufficiency for conducting high-quality SRs in the most influential journals of the field, which in turn might have led to the possible dissemination of inflated results. We strongly encourage the editors of the journals to take the issue of PB seriously and demand authors take measures to reduce its risk and use appropriate methods to assess its possible presence.

## Contributions

FM: conduct, analysis, and presentation of the research SB: design and presentation of the research

SI: design, conduct, and presentation

## Data Availability

All data produced in the present work are contained in the manuscript.

