## Appendix for "Publication bias in otorhinolaryngology meta-analyses in 2021"

### The title, journal, and type of the review of the 75 systematic reviews included in this study [1–75] are as follows:

| Journal | Title | Review type |
|---|---|---|
| Audiol. Neurootol. | Crucial Music Components Needed for Speech Perception Enhancement of Pediatric Cochlear Implant Users: A Systematic Review and Meta-Analysis | Effectiveness |
| Ear Hear | Occupational Hearing Loss Associated With Non-Gaussian Noise: A Systematic Review and Meta-analysis | Etiology and/or Risk |
| Ear Hear | Prevalence of Decreased Sound Tolerance (Hyperacusis) in Individuals With Autism Spectrum Disorder | Prevalence and/or Incidence |
| Int Forum Allergy Rhinol. | Effects of olfactory training on posttraumatic olfactory dysfunction: a systematic review and meta-analysis | Effectiveness |
| Int Forum Allergy Rhinol. | Effects of implants with steroids after endoscopic sinus surgery: A systematic review with meta-analysis of randomized controlled trials | Effectiveness |
| Int Forum Allergy Rhinol. | Usefulness of intraoperative frozen section for diagnosing acute invasive fungal rhinosinusitis: A systematic review and meta-analysis | Effectiveness |
| Int Forum Allergy Rhinol. | Topical nasal treatment efficacy on adult obstructive sleep apnea severity: a systematic review and meta-analysis | Diagnostic Test Accuracy |
| Int Forum Allergy Rhinol. | Botulinum toxin for chronic rhinitis: A systematic review and meta-analysis | Etiology and/or Risk |
| Int Forum Allergy Rhinol. | HPV in the malignant transformation of sinonasal inverted papillomas: A meta-analysis | Effectiveness |
| Otolaryngol Head Neck Surg. | Complications After Soft Tissue With Plate vs Bony Mandibular Reconstruction: A Systematic Review and Meta-analysis | Etiology and/or Risk |
| Otolaryngol Head Neck Surg. | Prognostic Significance of Extranodal Extension in HPV-Mediated Oropharyngeal Carcinoma: A Systematic Review and Meta-analysis | Etiology and/or Risk |
| Otolaryngol Head Neck Surg. | Tracheotomy in COVID-19 Patients: A Systematic Review and Meta-analysis of Weaning, Decannulation, and Survival | Effectiveness |
| Otolaryngol Head Neck Surg. | Systematic Review of Second Primary Oropharyngeal Cancers in Patients With p16+ Oropharyngeal Cancer | Prevalence and/or Incidence |
| Otolaryngol Head Neck Surg. | Systematic Review and Meta-analysis of the Change in Pharyngeal Bacterial Cultures After Pediatric Tonsillectomy | Effectiveness |
| Otolaryngol Head Neck Surg. | Pectoralis Major Onlay vs Interpositional Reconstruction Fistulation After Salvage Total Laryngectomy: Systematic Review and Meta-analysis | Effectiveness |
| Otolaryngol Head Neck Surg. | Meta-analysis Exploring Sinopulmonary Outcomes of Aspirin Desensitization in Aspirin-Exacerbated Respiratory Disease | Effectiveness |
| Otolaryngol Head Neck Surg. | Velcro Ties in Early Postoperative Pediatric Tracheostomy Care: A Systematic Review and Meta-analysis | Etiology and/or Risk |
| Otolaryngol Head Neck Surg. | Idiopathic Sudden Sensorineural Hearing Loss in Children: A Systematic Review and Meta-analysis | Etiology and/or Risk |
| Otolaryngol Head Neck Surg. | Systematic Review and Meta-analysis of Endoscopic vs Microscopic Stapes Surgery for Stapes Fixation | Effectiveness |
| Otolaryngol Head Neck Surg. | High-Risk Human Papillomavirus–Related Oropharyngeal Squamous Cell Carcinoma Among Non-Indigenous and Indigenous Populations: A Systematic Review | Etiology and/or Risk |
| Otolaryngol Head Neck Surg. | Olfactory Training for Postviral Olfactory Dysfunction: Systematic Review and Meta-analysis | Prevalence and/or Incidence |
| Otolaryngol Head Neck Surg. | Systematic Review and Meta-analysis: Effectiveness of Corticosteroids in Treating Adults With Acute Vestibular Neuritis | Effectiveness |
| Otolaryngol Head Neck Surg. | Antibiotic Prophylaxis for Thyroid and Parathyroid Surgery: A Systematic Review and Meta-analysis | Effectiveness |
| Otolaryngol Head Neck Surg. | Sinogenic Intracranial Suppuration in Children: Systematic Review and Meta-analysis | Etiology and/or Risk |
| Otolaryngol Head Neck Surg. | Factors Influencing the Development of Pneumonia in Patients With Head and Neck Cancer: A Meta-analysis | Effectiveness |
| Otolaryngol Head Neck Surg. | Prevalence and Characteristics of Taste Disorders in Cases of COVID-19: A Meta-analysis of 29,349 Patients | Effectiveness |
| Otolaryngol Head Neck Surg. | Surgical Management of Sialorrhea: A Systematic Review and Meta-analysis | Prognostic |

| | | |
|---|---|---|
| Otolaryngol Head Neck Surg. | Surgical Management of Bilateral Vocal Fold Paralysis in Children: A Systematic Review and Meta-analysis | Prognostic |
| Otolaryngol Head Neck Surg. | Management of Type 1 Laryngeal Clefts: A Systematic Review and Meta-analysis | Prevalence and/or Incidence |
| Otolaryngol Head Neck Surg. | Influence of Surgical Techniques on Endoscopic Dacryocystorhinostomy: A Systematic Review and Meta-analysis | Prevalence and/or Incidence |
| Otolaryngol Head Neck Surg. | Management of Flap Failure After Head and Neck Reconstruction: A Systematic Review and Meta-analysis | Effectiveness |
| Rhinology | Elective neck irradiation in the management of esthesioneuroblastoma: a systematic review and meta-analysis | Etiology and/or Risk |
| Rhinology | Intralymphatic immunotherapy for allergic rhinoconjunctivitis: a systematic review and meta-analysis | Effectiveness |
| Rhinology | Clinical effectiveness of house dust mite immunotherapy in mono- versus poly-sensitised patients with allergic rhinitis: a systematic review and meta-analysis | Prognostic |
| Rhinology | Anxiety and depression risk in patients with allergic rhinitis: a systematic review and meta-analysis | Effectiveness |
| Rhinology | Leukotriene receptor antagonist addition to intranasal steroid: systematic review and meta-analysis | Effectiveness |
| Rhinology | Is postoperative nasal packing after septoplasty safe? A systematic review and meta-analysis of randomized controlled studies | Etiology and/or Risk |
| Rhinology | Omalizumab for the treatment of allergic rhinitis: a systematic review and meta-analysis | Effectiveness |
| Laryngoscope | Tracheal Resection in the Management of Thyroid Cancer: An Evidence-Based Approach | Effectiveness |
| Laryngoscope | Autofluorescence and Indocyanine Green in Thyroid Surgery: A Systematic Review and Meta-Analysis | Prevalence and/or Incidence |
| Laryngoscope | Treatment of Vestibular Migraine: A Systematic Review and Meta-analysis | Effectiveness |
| Laryngoscope | Endoscopy-Assisted Transoral Approach to Resect Parapharyngeal Space Tumors: A Systematic Review and Meta-Analysis | Etiology and/or Risk |
| Laryngoscope | Outcomes of Adenotonsillectomy for Obstructive Sleep Apnea in Prader-Willi Syndrome: Systematic Review and Meta-analysis | Effectiveness |
| Laryngoscope | Sialendoscopy and Sjogren's Disease: A Systematic Review | Effectiveness |
| Laryngoscope | The Relationship between Croup and Gastroesophageal Reflux: A Systematic Review and Meta-Analysis | Etiology and/or Risk |
| Laryngoscope | Different Surgical Strategies in the Prevention of Frey Syndrome: A Systematic Review and Meta-analysis | Etiology and/or Risk |
| Laryngoscope | Association Between Human Papilloma Virus Infection and Malignant Sinonasal Inverted Papilloma | Prevalence and/or Incidence |
| Laryngoscope | Neuromodulators for Atypical Facial Pain and Neuralgias: A Systematic Review and Meta-Analysis | Effectiveness |
| Laryngoscope | Risk Factors for Multiple Tympanostomy Tube Placements in Children: Systematic Review and Meta-Analysis | Effectiveness |
| Laryngoscope | Lipoinjection for Unilateral Vocal Fold Paralysis Treatment: A Systematic Review and Meta-Analysis | Etiology and/or Risk |
| Laryngoscope | Pharmacological Treatments of Bell's Palsy in Adults: A Systematic Review and Network Meta-Analysis | Etiology and/or Risk |
| Laryngoscope | Scalar Translocation Comparison Between Lateral Wall and Perimodiolar Cochlear Implant Arrays - A Meta-Analysis | Etiology and/or Risk |
| Laryngoscope | Effect of Sleep Surgery on C-Reactive Protein Levels in Adults With Obstructive Sleep Apnea: A Meta-Analysis | Etiology and/or Risk |
| Laryngoscope | Method of Lateral Osteotomy to Reduce Eyelid Edema and Ecchymosis After Rhinoplasty: A Meta-analysis | Effectiveness |
| Laryngoscope | Usefulness of Sentinel Lymph Node Biopsy for Oral Cancer: A Systematic Review and Meta-Analysis | Etiology and/or Risk |
| Laryngoscope | Risk of Stroke and Myocardial Infarction After Sudden Sensorineural Hearing Loss: A Meta-Analysis | Etiology and/or Risk |
| Laryngoscope | Survival of Young Versus Old Patients With Oral Cavity Squamous Cell Carcinoma: A Meta-Analysis | Prognostic |
| Laryngoscope | A Meta-Analysis of 67 Studies with Presenting Symptoms and Laboratory Tests of COVID-19 Patients | Effectiveness |
| Laryngoscope | Nasal Peak Inspiratory Flow in Healthy and Obstructed Patients: Systematic Review and Meta-Analysis | Etiology and/or Risk |

| | | |
|---|---|---|
| Laryngoscope | Three-Dimensional Endoscopic Endonasal Surgery: A Systematic Review | Prevalence and/or Incidence |
| Laryngoscope | Barbed Reposition Pharyngoplasty versus Expansion Sphincter Pharyngoplasty: A Meta-Analysis | Effectiveness |
| Laryngoscope | Misperception of Visual Vertical in Peripheral Vestibular Disorders. A Systematic Review With Meta-Analysis | Effectiveness |
| Laryngoscope | Prevalence of Olfactory Dysfunction in Coronavirus Disease 2019 (COVID-19): A Meta-analysis of 27,492 Patients | Effectiveness |
| Laryngoscope | Paper Patching Versus Watchful Waiting of Traumatic Tympanic Membrane Perforations: A Meta-Analysis | Effectiveness |
| Laryngoscope | Residual Perforation Risk Assessment of Intratympanic Steroids via Tympanostomy Tube Versus Transtympanic Injections | Etiology and/or Risk |
| Laryngoscope | Complications of Neck Drains in Thyroidectomies: A Systematic Review and Meta-Analysis | Effectiveness |
| Laryngoscope | Bleeding Complications After Transoral Robotic Surgery: A Meta-Analysis and Systematic Review | Effectiveness |
| Laryngoscope | Prevalence of Sensorineural Hearing Loss in Pediatric Patients with Sickle Cell Disease: A Meta-analysis | Etiology and/or Risk |
| Laryngoscope | A Systematic Review and Meta-Analysis: Timing of Elective Removal of Tympanostomy Tubes | Etiology and/or Risk |
| Laryngoscope | Cochlear Implantation in Meniere's Disease: A Systematic Review and Meta-Analysis | Diagnostic Test Accuracy |
| Laryngoscope | Radiologically Defined Sarcopenia Affects Survival in Head and Neck Cancer: A Meta-Analysis | Prevalence and/or Incidence |
| Laryngoscope | Sudden Sensorineural Hearing Loss in Children—Management and Outcomes: A Meta-analysis | Effectiveness |
| Laryngoscope | Slide Tracheoplasty for Congenital Tracheal Stenosis Repair: A Systematic Review and Meta-Analysis | Etiology and/or Risk |
| Laryngoscope | A Systematic Review and Meta-Analysis of Taste Dysfunction in Chronic Rhinosinusitis | Prognostic |
| Laryngoscope | Effect of Endoscope Sinus Surgery on Pulmonary Function in Cystic Fibrosis Patients: A Meta-Analysis | Prevalence and/or Incidence |
